# The impact of substance use on tuberculosis treatment adherence among tuberculosis patients in Limpopo Province, South Africa

**DOI:** 10.1101/2023.06.08.23291161

**Authors:** Hulisani Matakanye, Takalani Tshitangano

## Abstract

**Background:** Substance use are associated with high discontinuation of tuberculosis treatment which is a barrier to tuberculosis control. Treatment discontinuation significantly increase the risk of poor treatment outcomes in both drug-susceptible and multidrug-resistant tuberculosis patients. This study aimed to explore the impact of substance use on tuberculosis treatment adherence among tuberculosis patients in Limpopo Province, South Africa.

**Method:** The study employed exploratory qualitative research design. Purposive sampling method was used to sample eight TB focal person and eight facility operational managers who were working in eight community health center within three selected districts in Limpopo province. Data were collected through key informant in-depth interviews and were analysed using Colaizzi’s method.

**Results:** The study results show four individual major themes that emerged from data analysis: (1) Patients forget to take treatment, (2) Patients miss follow up visit and DOT support appointments (3) Patients refuse social support (4) Patients refuse to attend health education and counselling.

**Conclusion:** The study highlighted serious impact of substance abuse on TB treatment adherence among TB patients. There is a need to integrate treatment for alcoholism and illicit drug users into primary health care to identify specific group of patients suffering from alcoholism and drug addiction on time, as a component of comprehensive TB treatment strategy.

## Introduction

Substance abuse is a major and growing devastating problem globally [1]. The increased usage of substance abuse has been seen to be high among younger people between the age of 15-30 years than that among older people and was shown by the Global Burden of Disease [2]. Younger people are also more likely to die from substance use. In 2019, alcohol use was found to be the leading risk factor for attributable burden of disease among people between the ages of 25 to 49, the second-leading risk factor among people between the ages of 10 to 24, and also the ninth-leading risk factor among all ages where it accounted for about 2.07 million deaths of males and 374,000 deaths of females, globally [3].

South Africa has emerged as one of the most attractive largest markets for illicit drugs in sub-Saharan Africa. Where the increased use of substance abuse has been noticed since 1994 as there is an ongoing increased demand in the promotion and availability of alcohol and other substances [4]. A relatively new cocktail psychoactive drug, known as nyaope, has flooded the drug market in South Africa. Cocktail drug nyaope is a street drug which is commonly found in South Africa, and is a mixture of low-grade heroin, cannabis products, antiretroviral drugs and other materials added as cutting agents [5]. The abuse of the cocktail drug nyaope in South Africa, has increased in over the years mainly amongst young people from poor backgrounds. Cocktail drug nyaope has become more popular between 2000 and 2006, in different provinces in South Africa. It is a very addictive cocktail of drugs and is commonly used in poor black and coloured townships in various provinces [5]. The problem of substance use is that they result in adverse health outcomes, and they are key drivers of poor TB treatment response amongst patients.

Literature shows that chronic substance abuse is associated with significant risk for medication non-adherence among patients with TB [6]. Heavy alcohol consumption impacts patient retention in care and is associated with an increased odds of loss to follow up [7]. One study shows that alcohol addiction is a risk factor for poor TB outcomes, treatment failure which sometimes result to death [8]. The misuse of both alcohol and illicit drugs during treatment is associated with discontinuing treatment, and these substances have common mechanisms to affect TB treatment outcomes, possibly leading to either a saturation or an amplification of their effects [9]. One study conducted in Limpopo province, South Africa found that, substance use among young and old people is one of the most signi?cant public health and social problems [10].

### Aim of the study

This study aimed to explore the impact of substance use on tuberculosis treatment adherence among tuberculosis patients in Limpopo Province, South Africa.

### Research questions

To better understands the impact of substance use on TB treatment adherence, the researchers investigate the following three nested sub-questions:

RQ1: What are the substances used by TB patients who are on TB treatment? RQ2: What are the effects of substance use on TB treatment adherence?

RQ3: What are the influences of substance use on the relationship of TB patients and health care workers and family members?

## Subjects and Method

### Study Design

The study used exploratory qualitative design to explore the impact of substance abuse on TB treatment adherence amongst TB patients. A qualitative approach of an exploratory nature was chosen because exploratory studies are qualitative in nature and perceptions are often explored. They are used to gain an understanding of underlying reasons, opinions, and motivations. They also provide insights into the problem. Exploratory designed allowed the researcher an opportunity to consult the stakeholders who shared their perceptions regarding the impact of substance abuse on TB treatment adherence.

### Study Site

The study was conducted in the Limpopo Province in the three selected districts of Waterberg, Capricorn, and Vhembe between 2020 and 2021. The researcher selected this study setting since there is dearth in data regarding the impact of substance abuse on TB treatment adherence among TB patients. The researcher selected eight community health center within three selected districts. Approximately 80% of the population in Limpopo province live in rural areas. Research indicates that there is a high substance use among people living in the rural areas.

### Study Population

The study population comprised of facility managers and TB focal person who are responsible for all TB related activities at the health care facility within eight selected health care facilities.

### Sampling Procedure

#### Sampling of districts

The sampling of the districts was done using a purposive sampling method to cover both rural and urban based districts of the province. Three districts; Vhembe, Capricorn and Waterberg district in Limpopo Province, the researcher intended to include two districts with lower TB treatment success rate and one district with higher TB treatment success rate, to understand and compare unique problems impacting those areas. Those three districts were chosen as they best served the purpose of the study.

#### Sampling of facilities

The sampling of the health care facilities was done using the non-probability purposive sampling technique. The researcher sampled a minimum of two and a maximum of four health care facilities in each selected district. Where the district has only two health care facilities then all of them were sampled. It was believed that participants found at these health care facilities would be able to expose impact of alcohol and illicit drug use affecting TB treatment adherence.

#### Sampling of participants

Purposive sampling method was used to sample site TB focal person, and facility operational manager who were readily available at the time of data collection and were willing to participate in the study [11]. Researcher recruited those participants because it was believed that they possessed quality of the selected information to explore impact of substance abuse on TB treatment adherence.

### Inclusion and Exclusion Criteria

Only TB focal persons from the age of 18 years old who had more than three years’ work experiences have been included in the study, and those with less than three years’ experience were not included. Facility operation managers who were responsible for TB program and the operation of the facility were included in the study. Facility managers who were not responsible for TB program were not included in the study.

### Sample Size Determination

Qualitative data are small enough to allow the deep, case-oriented analysis, therefore fewer participants are needed [12]. For key informant interview, eight TB focal person (one from each facility), and eight TB facility operational managers (one from each facility), with quality of the selected information who were readily available were identified in this study.

### Data Collection Procedure

Data was collected through in-depth individual interviews. All interviews were conducted in English. Researcher used face-to-face interactions as a primary method to recruit participants at the health care facilities. All participants who signed a written informed consent form before data collection were included in this study and were interviewed. The researcher explained the purpose of the study to each participant before interviewing them. Participants were assured that confidentiality will be maintained. The interview dates and times were arranged with participants prior to data collection date. Data collection took place between June 2020– September 2020. The interviews were conducted early in the morning as suggested by participants before they could start with their duties to avoid distracting them from their normal ward routine work. Separate interviews with 16 adults’ participants (14 females and 2 males) took place in the private offices, and each lasted approximately 30 minutes. All interviews were conducted by the researcher to ensure the quality of the data. Questions were asked per identified factors; and was deliberated on as long as the participant could narrate.

The research participants freely responded to open-ended questions in narrative form using their own words, thus sharing their own perspectives with the researcher. Questions were not planned in an inflexible manner. The questions were not asked in a pre-arranged sequence, but the researcher ensured that all relevant topics were covered and that the research focus was kept in mind. The researcher also asked probing questions to guide participants to elaborate further upon their responses where additional information were required or where unclear answers required more clarity. This resulted in gaining in-depth accounts about the impact of substance use on TB treatment adherence. Data were collected by means of audio-recordings, field notes, and in-depth interviews.

### Data Management and Analysis

Data was stored in a password-protected computer. Access to the database was restricted to the researcher and supervisors only. Data were stored as per university’s protocols. Any identifiable information that was collected remained confidential and was only accessible to the researcher and supervisors, however, participants were not asked their names or any personal information to maintain anonymity. Data were analysed in groups not individually to avoid identifying the participants by their responses. Qualitative data analysis always takes place concurrently with data collection. Therefore, the researcher attempted to gather, manage, and interpret a growing bulk of data simultaneously. In this study, the audio-recorded interviews were transcribed and coded immediately after data collection.

Data were analysed using Colaizzi’s [13] methods. In this study each research participant’s verbatim transcript was read to acquire a sense of the whole. Significant statements and phrases pertaining to the phenomenon being studied were extracted from each transcript. Meanings were formulated from the significant statements. Meanings were organized into themes, and themes evolved into theme clusters and eventually into theme categories. The results were integrated into a rich and exhaustive description of the lived experience. Validation was sought from the research participants to compare the researcher results with their lived experiences.

### Ethical Consideration

The proposal was submitted and presented to the School of Health Science and the University Higher Degrees Committee (UHDC), and an ethical clearance was granted (SHS/19/PH/28/0411). Permission to conduct the study was obtained from the Limpopo Provincial Department of Health and Vhembe District, Waterberg District, and Capricorn District Department of Health. Furthermore, permission was obtained from the facilities’ operational managers.

Researcher used face-to-face interactions as a primary method to recruit participants. All participants who were willing to participate in the study were asked to complete individual written informed consent forms. The nature of the research was described to each participant, and they were all informed of their right to refuse to participate, or to withdraw from participating if they felt that they could not continue. Participants were also informed and assured that the information within their responses would not be used against them or shared with other third person but would only be reported as study findings.

Anonymity was also ensured as participants did not write down their names or any personal identification. The researcher respected the choices and agreements made with the participants. Individuals were not victimized for refusing to participate.

## Results

After data organization and analysis, five individual major themes were formulated from the findings of this study (See figure 1). Participants shared their perceptions regarding the impact of alcohol and illicit drug abuse on TB treatment adherence in Limpopo Province.

**Figure 1:**
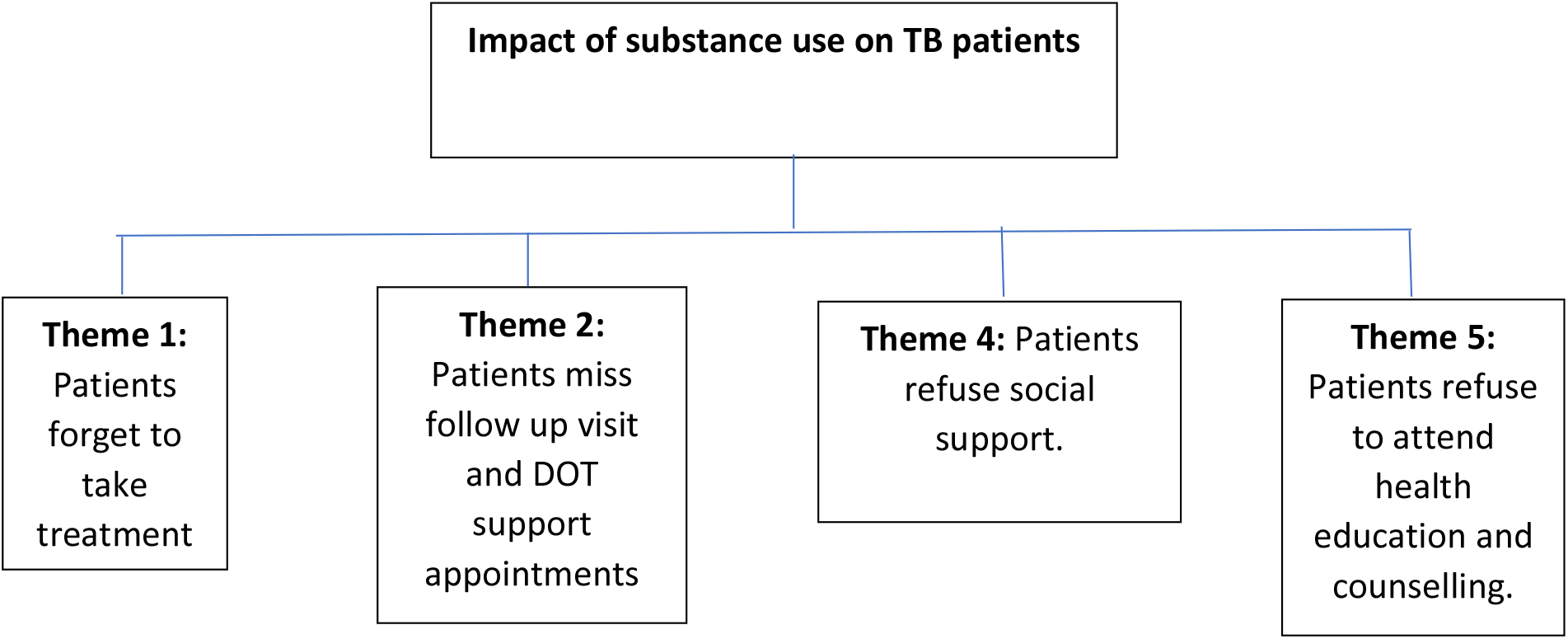
Themes for the impact of substance use on tuberculosis treatment adherence among TB Patients.

Four main themes emerged after extensive data organization and analysis on the issue of alcohol and illicit drug abuse. These individual themes were developed from the comments that were made by the participants during data collection. A discussion of findings follows the presentation of each theme, as it is asserted that integrating the findings and the discussion is an appropriate method for encapsulating the essence of the phenomenon under investigation. Existing literature was searched and used to support the findings of this study. Participants revealed that the substances that are used by TB patients are nyaope (which is a South African street cocktail drug, mixture of low-grade heroin smoked with cannabis) and alcohol.

### Theme 1: Patients forget to take treatment

Participants revealed that TB patients who abuse both alcohol and cocktail drug nyaope have been observed to have a problem of always forgetting to take their treatment. Participants further indicated that, patients on substance abuse spend too much time at the shebeens and on the street, they do not have time to take their treatment at all. Health care workers indicated that, they find it hard to monitor those patients while taking their TB treatment.

One TB focal person (Participant 11 – female) indicated that: “…*these patients are always drunk, and they are always at the shebeens and the way they are, it looks like they are trying to numb the pain*.

Another TB focal person (Participant 9 – female) added: *These patients do not have time to take their treatment. I think some of them get drunk to a point where they even forget that they are sick, and it is difficult to monitor them*.

### Theme 2: Patients miss follow up visit and Directly Observed Therapy (DOT) support appointments

Participants revealed that TB patients who use cocktail drug nyaope and alcohol do not honor their follow up visits or their DOT support appointments as most of them are always not at home. Even if DOT supporters follow them at home or their usual spots, they do not find them. If they find them, they run away the moment they see DOT supporters, because they do not want their friends to know that they are sick. The problem is those patients do not disclose their status as they fear to be discriminated or isolated. They end up spreading the disease in the communities as they spend time in overcrowded areas or poorly ventilated homes or social venues.

One facility operational manager (participant 13 – female) said: *“Bo-nyaope (mean people taking illicit drug called nyaope) don’t want to see our DOT supporters they will agree to have home-based carers to support them, but the moment they see them they run away, they say that their friends will laugh at them and call them weak”*.

Another TB focal person (Participant 15 - male) added that: *“Some of them (patients) we do not even get to see them because they wake up very early in the morning and go to shebeens and they come home very late at night, and they do not even communicate with home-based care givers”, and they do not bother to go for follow up visit*.

### Theme 3: Patients refuse social support

Social support from family, friends and healthcare workers plays an important role for patients on treatment as it helps them adhere to their treatment. Patients who are supported feel encouraged to take their treatment. However, this study shows that TB patients who are on substance abuse refuse social support as they fear to be reprimanded for their behaviour. Most patients do not even bother to disclose their TB status and as a result they are not supported.

One facility operational manager (participant 10 – female) indicated that: *these nyaope boys refuse support even from us or their family. Some are staying on the street, and they move from one place to another, the moment we know where they*.

Another TB focal person (participant 5 – male). *Those nyaope boys do not disclose their status and if their family find out that they are sick, they will rather leave home so no one can reprimand them to stop using nyaope*.

### Theme 4: Patients refuse to attend health education and counselling

TB treatment non-adherence often results from inadequate knowledge or understanding of the disease and its treatment. Adherence to the long course of TB treatment is a complex process that requires better understanding of the disease and the treatment. During the interview’s participants revealed that patients on substance abuse do not want to attend health education and counselling. They are always in a hurry to leave the healthcare facility and if healthcare workers are taking long or if the queue is too long, they complain and leave. Some patients only visit the facility whenever they are feeling too sick.

One facility operational manager (participant 6 – female) stated that: *“…Another challenge that we have is that some patients are mostly ignorant, they do not want any counselling and health information, they only want to collect their treatment and leave. Most of the nyaope people are very inpatient*.

Another TB focal person (participant 14 – male) indicated that: *“It is too difficult to educate them, they don’t listen because they are in a hurry to leave. sometimes we ask if they can come with someone from their family who can better understand the information. It looks like they do not want their families to be involved as they know they will ask them to stop taking nyaope”*.

## Discussion

This study discovered that the substance that is used by most TB patients in the rural area in Limpopo province is alcohol, and cocktail drug nyaope. The study further revealed that the use of those substances causes those TB patients to forget to take their treatment which in turn makes them to default their treatment and has also have huge impact on their treatment outcome. Patients were found to be spending too much time at the shebeens and they don’t have time to take their treatment, and as a result majority of them always forget to take their treatment. Other study shows that among patients with TB who interrupted the course of treatment, about 47.7% were heavy alcohol drinkers [14]. According to the study that was done by Shruthi et al. [15], stated that elderly patients who abuse alcohol were not be fully compliant to a long-term medication. Alcohol abuse has also been associated with forgetting to take treatment (in 7.5% cases) and consequently defaulting [16].

The study further showed that TB patients who are suffering from substance use do not honour their DOT appointments or follow up visit. One study shows that heavy alcohol use impacts retention in care and is associated with missed follow up visits as it is not easy to monitor such patients [7]. Another study shows that heavy substance use is associated with missed DOT visits, where most MDR TB patients who consumed alcohol during treatment have missed an average of 18 more intensive phase doses [17].

The findings of this study further revealed that majority of patients suffering from substance use are afraid to disclose their status and refuse social support when taking treatment as they are afraid to be reprimanded and stigmatized. Participants alluded that family support is important for TB patient to adhere to their treatment especially during the intensive phase, however, according to the literature, stigma make patients afraid to ask for social support, thereby reducing adherence [18].

This study shows that patients suffering from substance abuse refuse to attend health education and counselling as they are always in a hurry to leave the health care facility. One study shows that, being aware of the potential risk of non-adherence among patients on alcohol and substance abuse may enable the healthcare workers to undertake additional educational efforts with those patients to emphasize the importance of treatment adherence [19]. Literature shows that patients’ understanding of the disease and treatment including the duration required to take treatment and the consequences of defaulting has influence on adherence [20].

The findings of this study disclosed that substance abuse such as cocktail drug nyaope and alcohol among TB patients have a huge impact treatment adherence. it is important to provide additional health education to patients on substance abuse for them to understand the impact substance abuse on treatment outcome.

### Limitations

The study focused on the perception of the health care workers’ regarding the impact of illicit drugs and alcohol abuse on tuberculosis treatment adherence amongst tuberculosis Patients in Limpopo Province, South Africa and therefore the findings cannot be generalized. However, according to Smith [21], generalizability in a qualitative study is not intended. The study was conducted in a predominately rural province of South Africa. It is likely that if urban areas were included, this could have led to different data findings. As a qualitative study, the results are not generalizable to other settings or populations. However, they do give an insight into this setting and aim to further understand the impact of substance abuse on TB treatment adherence among TB patients who are suffering from alcohol and drug abuse.

## Conclusions

Although the study involved a small sample group, the findings provide important insights and highlighted that substance abuse are key mechanisms of TB treatment non-adherence. Based on the findings of the study, there is a need to integrate treatment for substance use addiction into primary health care and identify specific patients suffering from alcoholism and drug addiction on time. The study shows that failure to do so among TB patients suffering from both diseases shows a missed opportunity with serious clinical and public health implications. It is important to implement patient’ psychological assessments, to screen patients’ substance use disorder to identify patients who need help on time. Further studies are needs to assess the mental health status of TB patients with substance use disorder to determine appropriate interventions and also develop strategies to manage psychiatric complications in TB patients in order to improve treatment adhere. There is also an urgent need to conduct continuous community engagement by health care workers who have participated in the study to raise awareness on the impact of substance abuse on TB patients who are on treatment.

## Data Availability

Authors do not have permission to share raw data publicly to maintain confidentiality. Permission to collect data was granted by Provincial Department of health and the researcher agreed not to share raw data with any third person as they were meant specifically for the research project. Participants signed written informed consent form and were assured that data will be confidential and will only be used for the purpose of the study. Analyzed data is available from the researcher with meaningful request.

## Recommendations

Based on the above findings, the following recommendations are made:

- There is a need to integrate treatment for patients on substance abuse into primary health care to identify specific group of patients suffering from alcoholism and drug addiction on time, as a component of comprehensive TB treatment strategy.
- It is important to design health education in a way that it will encourage patients to disclose their status to their family members so they can receive proper support.
- The Department of Education should collaborate with Department of Health to intensify the implementation of school-based alcohol and drug prevention programmes that will help reduce alcohol and drug use among youth.

## Acknowledgments

The authors wish to thank the University of Venda and Limpopo Province Department of Health, for granting ethical clearance and permission to conduct the study. The participants are also thanked for their consent to participate in the study. The supervisors are thanked for their tireless efforts and guidance.

## Author Contributions

H.M. was the project leader and involved in the research instrument development, data collection, data analysis, and wrote the article. T.G.T., was the supervisor of the study and was also responsible for supervising the writing of this article. All authors have read and agreed to the published version of the manuscript.

## Informed Consent Statement

Written informed consent was obtained from all subjects involved in the study.

## Conflicts of Interest

The authors declare no conflict of interest.

## Notes

### Competing Interest Statement

The authors have declared no competing interest.

### Funding Statement

The authors received no specific funding for this work.

### Author Declarations

The proposal was submitted and presented to the School of Health Science and the University Higher Degrees Committee (UHDC), and an ethical clearance was granted (SHS/19/PH/28/0411).

